# Segmentation-Free Pretherapeutic Assessment of BRAF-Status in Pediatric Low-Grade Gliomas

**DOI:** 10.1101/2025.01.30.25321339

**Authors:** Kareem Kudus, Matthias Wagner, Min Sheng, Julie Bennett, Anthony Liu, Uri Tabori, Cynthia Hawkins, Birgit Betina Ertl-Wagner, Farzad Khalvati

## Abstract

**Background:** BRAF status is crucial for treating pediatric low-grade gliomas (pLGG) and can be assessed non-invasively from segmented tumor regions on MRI using machine learning (ML). However, there are inherent limitations to manual and automated tumor segmentations.

**Purpose:** To assess the performance of automated segmentation algorithms and to develop and assess a segmentation-free ML classification pipeline that identifies BRAF status from whole-brain FLAIR MRI sequences.

**Materials and Methods:** In this REB-approved retrospective study, molecularly-characterized tumors and whole-brain FLAIR MR images were collected from 455 patients with pLGG treated between 1999 and 2023 at a single tertiary care children’s hospital. We trained and evaluated three medical segmentation models, TransBTS, MedNeXt, and MedicalNet. Next, we developed a model to identify BRAF status from whole-brain FLAIR MRI, without any reliance on manual or automated segmentations. We then implemented a novel pretraining regimen that embedded segmentation knowledge into the whole-brain FLAIR MRI classification model. Finally, we trained and evaluated a baseline model that used manual segmentations as inputs. All ML models were trained and evaluated under a nested-cross validation scheme, and mean performance across all test folds was compared using the corresponding t-test.

**Results:** The MedNeXt segmentation model (mean Dice score: 0.555) outperformed both the convolutional neural network (CNN) based MedicalNet (0.516) and the CNN-transformer hybrid TransBTS (0.449) (p <0.05 for all comparisons). The MedNeXt style classification model achieved a one-vs-rest area under the ROC curve of 0.741 using the whole brain FLAIR sequence as an input, without any segmentation knowledge. This was improved to 0.772 through pretraining on the segmentation task, which was not significantly different from the baseline manual segmentation-based model (0.756, p-value: 0.141).

**Conclusion:** BRAF status can be assessed non-invasively using ML models based on whole-brain FLAIR sequences. Dependence on inconsistent manual or automated segmentations can be reduced by integrating tumor region information into the model through pretraining.

## Introduction

Pediatric low-grade gliomas (pLGG) are the most common brain tumor in children, accounting for approximately 40% of pediatric central nervous system (CNS) tumors ^1^. As a diverse set of tumors, pLGGs can occur throughout the CNS and consist of various different histopathologies ^2^. Despite the heterogeneity of pLGGs, it has been found that these tumors are often the result of a select few molecular alterations in the mitogen-activated protein kinase pathway, most commonly KIAA1549::BRAF (BRAF Fusion) or point mutation of BRAF p.V600E (BRAF Mutation) ^3^. BRAF status has proven to be crucial for accurate prognostication and risk assessment of pLGG and for therapeutic decision-making ^4,5^. The importance of genetic status is highlighted in recent editions of the WHO CNS tumor classification ^6^. While historically CNS tumor classification has been based primarily on histological findings, the more recent approaches also rely on molecular status ^7^.

The identification of the molecular alterations driving pLGGs has been transformative for their treatment. Therapeutics targeting specific genetic alterations that can supplement or replace classic cytotoxic treatments have been developed ^4,8^. Usage of these targeted therapies, optimal prognostication, and risk assessment require surgically acquired tumor tissue which is used to determine BRAF status ^9^. Currently, in cases where tissue samples cannot be retrieved, such as with difficult-to-access tumors, the disease cannot be well characterized, and it is difficult to identify an appropriate targeted therapy.

Machine learning (ML) has shown potential as an alternative, non-invasive method of detecting BRAF status in patients with pLGG. Earlier studies used ML to successfully classify patients with pLGG by genetic status based on manually segmented tumor regions of their MRIs ^10–16^. While encouraging, the pipelines described in these works depend on manual tumor segmentations that have known intra-reader ^17^ and inter-reader ^18^ variability issues. More recent studies have aimed to eliminate the reliance on manual segmentations by introducing more reproducible pipelines incorporating automated segmentation models ^19–21^. However, such an approach is not fully reliable either, given that earlier studies focused on automating the segmentation of pLGGs either excluded difficult and outlier cases ^22^, or failed for certain patients, missing the tumor entirely ^23^.

In this study, we propose a non-invasive BRAF status identification pipeline that aims to remedy the weaknesses of earlier studies that relied on manual or automated segmentations. Previous works attempting to automate the segmentation of pLGGs ^19,23^ used deep learning (DL) models based on the U-Net ^24^. These DL architectures, which rely on convolutional neural networks (CNN), were state-of-the-art until recently. Now, however, more advanced models, such as those relying fully or partially on transformer layers ^25,26^, or on the ConvNeXt ^27^ architecture, have proven more effective for adult brain tumor segmentation. We aimed to evaluate the performance of each of the aforementioned DL architectures for pLGG segmentation, in order to develop a more reliable automated segmentation model capable of consistently identifying the tumor region. We also aimed to develop and evaluate a novel ML pipeline to determine BRAF status from whole-brain fluid-attenuated inversion recovery (FLAIR) MRI sequences, relying on manual tumor segmentations, and features derived from them, for pretraining only, rather than using manually or automatically identified tumor regions as model inputs. Once deployed, this pipeline would have no reliance on tumor segmentations whatsoever.

## Materials & Methods

This study conformed to the guidelines and regulations of the research ethics board of our institution, which approved this retrospective study and waived the need for informed consent due to the retrospective nature of the study. All data were deidentified after being extracted from the electronic health record database.

### Data

A total of 513 patients treated for pLGGs located in the brain parenchyma from 1999 to 2023 at The Hospital for Sick Children (SickKids) (Toronto, Canada) were identified using the SickKids electronic health record database. We excluded 15 patients for which genetic information was unavailable. We excluded 43 additional patients who did not have a 2D axial or coronal FLAIR MR sequence available. The demographics of the 455 patients included in this study are summarized in Table 1.

**Table 1:** Demographic information of the patient population. Patients without BRAF alteration were tested and found to be negative for BRAF Fusion and Mutation. The class “Non-BRAF altered” included tumors where other more rare genetic alterations were identified (65 patients), and those where the underlying genetic driver of pLGG could not be identified (57 patients). The number and proportion of patients with a tumor located in the supratentorial region is listed, the remaining patients had tumors found in the infratentorial region. Pathologies identified through analysis of tumor samples are listed. Other pathologies included were Pilomyxoid Astrocytoma, Gangliocytoma, Oligodendroglioma, and additional less common pLGGs.

|  |  | By Class |  |  |
| --- | --- | --- | --- | --- |
|  | Overall | BRAF Fusion | BRAF Mutation | Non-BRAF Altered |
| **Number of patients** | 455 (100%) | 190 (41.7 %) | 95 (20.9 %) | 170 (37.4 %) |
| **Male (%)** | 54.3 | 50.0 | 56.8 | 57.6 |
| **Mean patient age (Years)** | 8.6 | 7.3 | 9.7 | 9.4 |
| **Tumor Location (Supratentorial)** | 282 (62.0%) | 59 (31.0%) | 84 (88.4%) | 139 (81.8%) |
| **Pathology** | - | - | - | - |
| **Pilocytic Astrocytoma** | 151 (33.2%) | 114 (60%) | 8 (8.4%) | 29 (17.1%) |
| **Low-Grade Astrocytoma** | 69 (15.2%) | 13 (6.8%) | 19 (20%) | 37 (21.8%) |
| **Ganglioglioma** | 45 (9.9%) | 7 (3.7%) | 24 (25.3%) | 14 (8.2%) |
| **Dysembrylastic Neuroepithelial Tumor** | 25 (5.5%) | 0 | 2 (2.1%) | 23 (13.5%) |
| **Diffuse Astrocytoma** | 22 (4.8%) | 3 (1.6%) | 9 (9.5%) | 10 (5.9%) |
| **Other** | 143 (31.4%) | 53 (27.9%) | 33 (34.7%) | 57 (33.5%) |

### Molecular Subtyping

We followed the same stepwise process for molecular characterization as previous works which relied on subsets of the data we used in this study ^11,13,16^. Formalin-fixed paraffin-embedded tissue obtained during biopsy or resection was used for most patients. If not available, frozen tissue was used. BRAF mutations were detected using immunohistochemistry, sequencing, or droplet digital PCR. BRAF fusions were identified using either fluorescence in situ hybridization or a nCounter Metabolic Pathways Panel (NanoString Technologies). Patients not found to be positive for BRAF mutation or fusion were determined to be “non-BRAF altered”.

### Image Acquisition, Segmentation and Preprocessing

Patients underwent brain MRI at field strengths of 1.5T or 3T using MRI scanners of different vendors (Signa, GE Healthcare; Achieva, Philips Healthcare; Magnetom Skyra, Siemens Healthineers). Tumor regions were segmented semiautomatically using the level tracing-effect tool of 3D Slicer ^28,29^ (Version 4.10.2) by a pediatric neuroradiology fellow (MS) and validated by a fellowship-trained pediatric neuroradiologist (MWW). Images were bias-corrected, resampled to a resolution of 240×240×155, z-score normalized and registered to the SRI24 atlas ^30^ using 3D Slicer.

### ML pipelines for pLGG genetic status classification

At a high level, we explored two approaches to designing a pipeline capable of classifying pLGGs into three classes: BRAF Fusion, BRAF Mutation, and non-BRAF altered, and did not rely on manual or automated segmentations. First, we assessed the potential for advanced DL models to improve the performance of automated segmentation models, making these segmentations more reliable. We then introduced a novel whole-brain FLAIR-based segmentation-free classification model. Figure 1 outlines the experiments that were used to assess both approaches, details of which can be found below.

**Figure 1:**
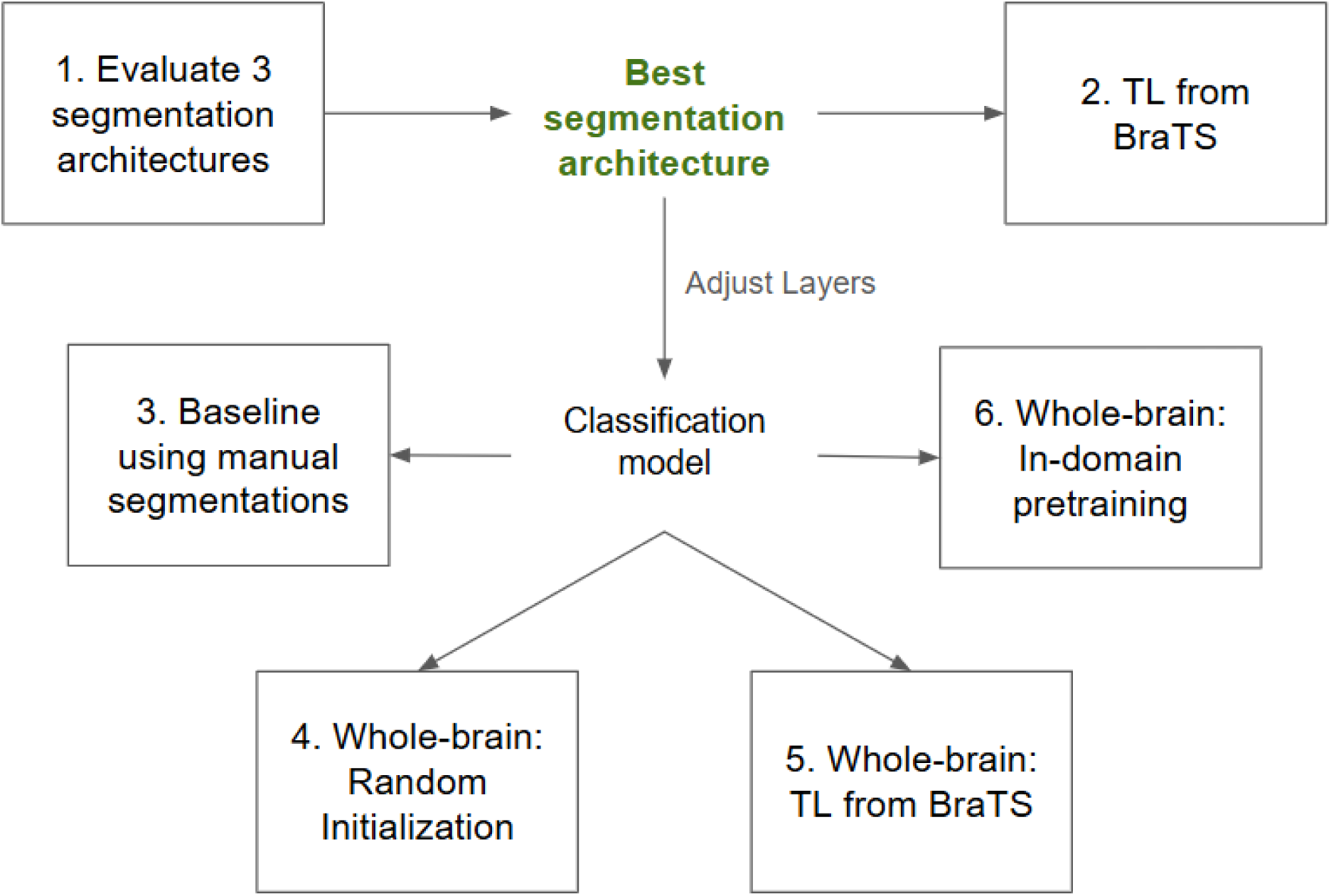
A high-level overview of our methodology. We started by testing 3 different architectures on the task of pLGG segmentation (1). The architecture that performed best on this task was used for all ensuing experiments. We then tested whether transfer learning from an adult brain tumor dataset could help improve segmentation performance (2). Next, the segmentation architecture was converted into a classification model by adding some layers and removing others. The classification model was trained and evaluated with manually segmented tumor regions as input (3) to generate baseline results. Finally, the classification model was trained and tested with whole-brain FLAIR MRIs as input using three different initialization schemes: random (4), transfer learning (5), and pretraining (6)

### Advanced DL models for more accurate tumor segmentation

We evaluated the performance of three different DL architectures for tumor segmentation on our pLGG dataset: 1. a CNN based on the MedicalNet architecture ^31^ 2. a CNN, transformer hybrid similar to TransBTS ^26^, and 3. a MedNeXt ^32^ style ConvNeXt-based ^27^ model. A review and comparison of these architectures is provided in the Appendix. We relied on the GitHub repository of the referenced publications where these architectures were introduced for implementation. The number and size of layers were adjusted to obtain a similar number of trainable parameters (approximately 400k) within each model for a fair comparison between architectures. We ran all subsequent experiments with the architecture that was found to perform best on this segmentation task because it showed the highest potential to derive insights from MRIs of pLGGs (Figure 1)

Next, we explored whether transfer learning (TL) from the Brain Tumor Segmentation (BraTS) challenge dataset [12], which consists of MRIs of adult patients with brain tumors, could improve the performance of our segmentation models (Figure 2). TL aims to improve performance on one task by leveraging knowledge learned from a related task in advance ^33^. It involves pretraining a model on a large dataset before fine-tuning it on a smaller target dataset with the model learning to extract useful features on the larger dataset. With this knowledge transferred to the smaller target dataset, the model is less likely to overfit compared to when training from scratch. To assess the effects of TL, the model was first pretrained on the BraTS dataset, where the provided FLAIR images and manual segmentations were used as model inputs and outputs respectively. Subsequently, the model was fine-tuned and evaluated on our pLGG dataset.

**Figure 2:**
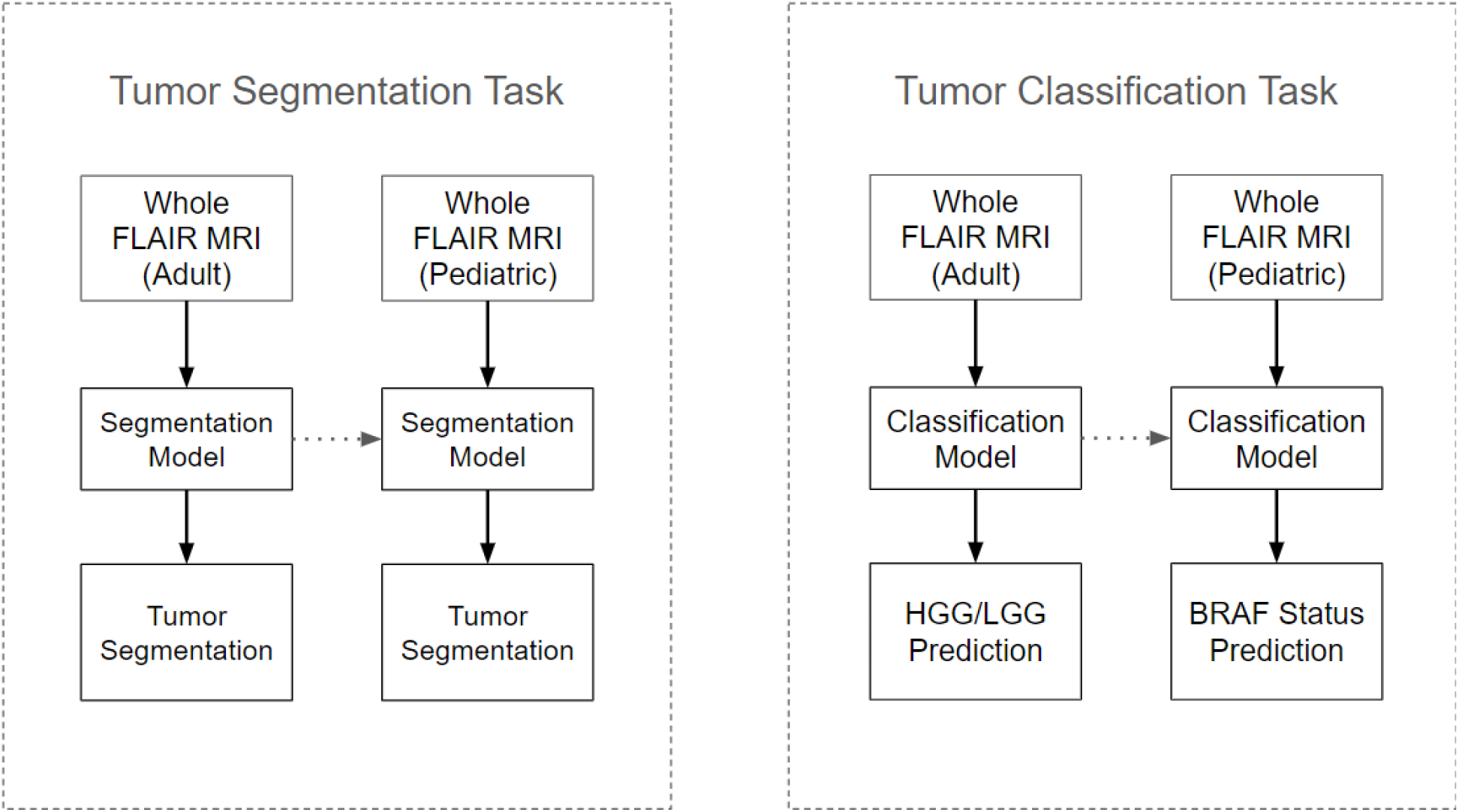
Depiction of BraTS TL regimen for the segmentation (left) and classification (right) tasks. The input to all models was the whole-brain FLAIR MRI from the respective dataset. The model was first trained on the BraTS dataset to identify tumor regions and HGG vs LGG, for the segmentation and classification tasks respectively. The pretrained weights were then transferred over and trained on the corresponding task using the pLGG data.

### Segmentation-free whole-brain FLAIR classification

The architecture that performed best for pLGG segmentation through the experiments detailed above was reconfigured for classification. The decoder was discarded, and a fully connected layer with three outputs, one for each of the three classes (BRAF Mutation, BRAF Fusion, and non-BRAF altered), was added on top of the encoder. The resultant model was randomly initialized and then trained to identify pLGG genetic subtype from whole-brain FLAIR MRI sequences, rather than using the tumor region alone. Next, similar to the experiments detailed above for segmentation, we attempted to improve classification model performance through transfer learning using the BraTS dataset (Figure 2). The model was first trained to differentiate between high-grade and low-grade gliomas using whole-brain FLAIR MRI sequences from the BraTS dataset, then fine-tuned on our BRAF status classification task.

In another attempt to improve classification performance, we implemented a pretraining regimen (Figure 3) that relied on a set of tasks we hypothesized would give the model a good understanding of the tumor region. The tasks were tumor segmentation, location identification, and radiomic feature value prediction, and the model was trained on them simultaneously using the sum of the three loss functions. The tumor segmentation task was the same as described in the first set of experiments (1). The location identification task required the model to predict whether the tumor was located in the supratentorial or infratentorial region of the brain. The label for this binary classification task was manually identified through analysis of the FLAIR MRI sequence by a pediatric neuroradiologist (MWW). Radiomics involves extracting quantitative features from medical images ^34^; it has been shown that these features are predictive of BRAF status ^11,13^. We extracted radiomic features from the manually defined tumor region and used them to train a BRAF status random forest prediction model for patients in the training and validation sets according to the open-radiomics protocol ^35^. The top 10 radiomics features were selected according to permutation importance; the values of these features were used for pretraining. If pretraining worked as expected, the model would recognize both where the tumor was located, and characteristics within the tumor that are correlated with BRAF status. As illustrated in Figure 3, after pretraining, the model was converted into a classification model, with certain pretrained layers kept intact, before the entire model was finetuned on the pLGG classification task.

**Figure 3:**
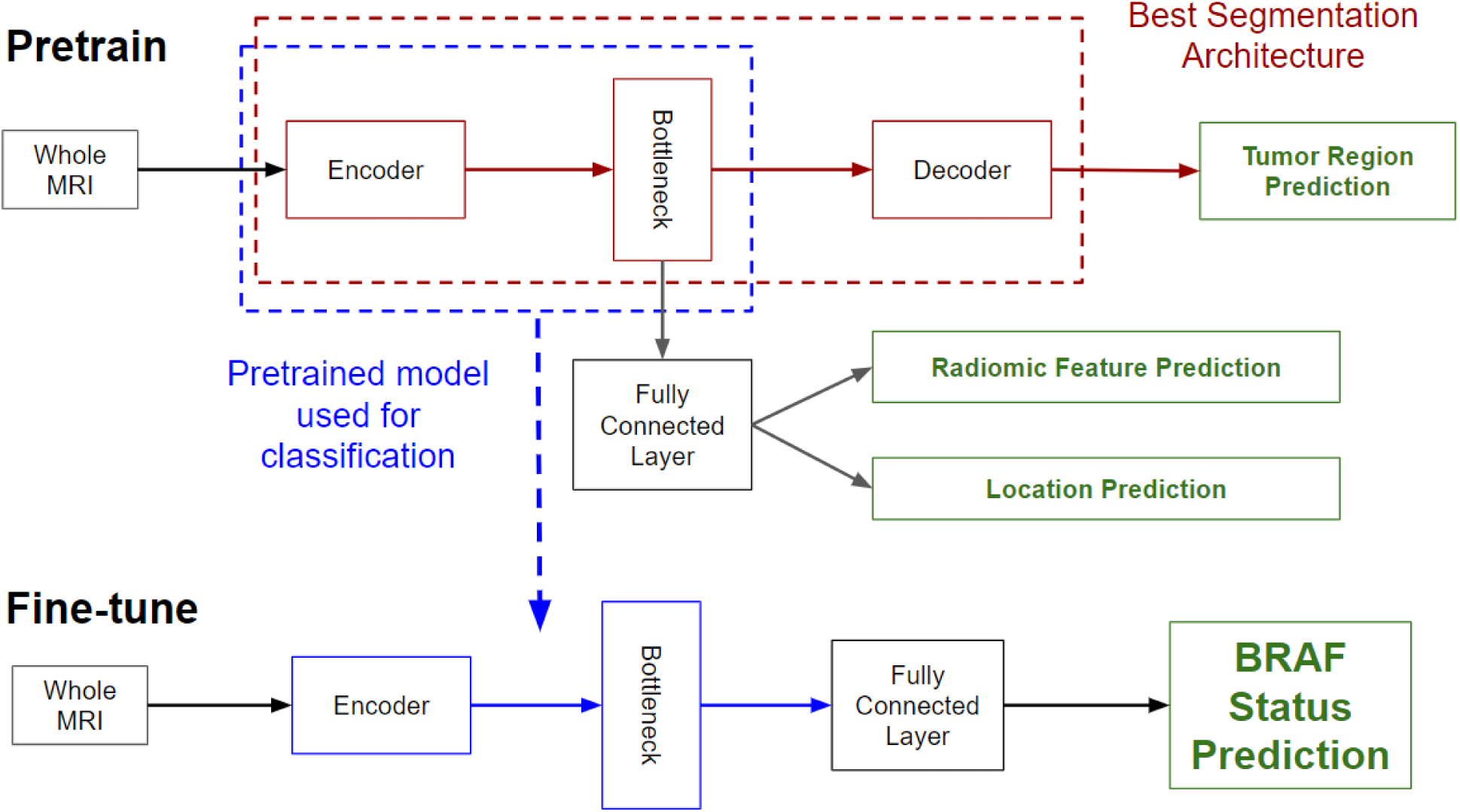
Overview of the pipeline for our best-performing BRAF status classification model, which uses manual tumor segmentations, radiomic feature values, and tumor locations for pretraining. The top of the image describes the pretraining phase, where the whole MRI is used as the input. In red is the architecture that was found to be best for tumor segmentation, which processes the image and outputs the predicted tumor region (top right, in green). During pretraining, a fully connected layer was added to the segmentation architecture. The output of the bottleneck flowed into the fully connected layer, which output radiomic feature values and tumor location (middle right, in green). The fully connected layer consisted of 11 neurons (10 radiomics features, one location feature). The bottom of the figure describes the fine-tuning phase, which relied on the pretrained weights in the blue box (encoder and bottleneck layers). These layers were transferred over directly from the pretrained model and used as the starting point for training the final BRAF status classification model. A new fully connected layer consisting of just three neurons (one for each class) was added, to take in the bottleneck output, and generate a BRAF status prediction (bottom right, in green). The same pipeline described at the bottom of the figure for fine-tuning was used for the experiment where we aimed to identify BRAF status from whole MRIs without pretraining, except the model parameters were randomly initialized.

### Classification based on tumor region alone

Most previous studies aiming to identify pLGG genetic status used manually segmented tumor regions as inputs to their ML models. Thus, a final set of experiments, which used manual segmentations alone as input to the same classification architecture described above, was run and served as a baseline.

### Experiment Configuration

We trained and evaluated all models using a nested-cross validation scheme, where patients were divided into train (80%), validation (10%), and test (10%) sets 25 times using a stratified split to ensure representative proportions of each class within each split. In each of these 25 trials, three separate models were trained on the training set using three different learning rates (0.00001, 0.0001, 0.001). Validation loss was calculated on the validation set at the end of each epoch. The optimal model selected for final evaluation on each trial was chosen according to the learning rate and epoch that resulted in the lowest validation loss. Reported performance metrics were based on the unseen test set, which was only used for model evaluation.

Kaiming Normal weight initialization was used to initialize all model parameters, where pretraining was not employed. Dropout was used to train all models (0.75 for fully connected layers, 0.25 for convolutional layers where entire channels were zeroed out) and turned off for interference. Cosine annealing was used to decay the learning rate over a maximum of 200 epochs for classification, and 50 for segmentation, which was slower. Training was stopped early to conserve resources if validation loss did not fall for 10 epochs in a row. The batch size was 8 images for classification experiments, but only 2 for segmentation experiments which required more memory. Gradient accumulation was used for the segmentation experiments, to effectively increase the batch size by only updating model parameters after every other batch.

Cross-entropy loss, weighted by the proportion of patients in each class, was used to train classification models. Classification models were primarily evaluated according to the One-Vs-Rest Area Under the Receiver Operating Characteristic Curve (AUC). Segmentation models were trained using the sum of soft dice loss and binary cross entropy loss and evaluated by the Dice score. During pretraining, the radiomic feature value prediction and location identification tasks were trained using a smooth L1 loss, and binary cross-entropy loss, respectively. All experiments for all models used identical data splits, allowing for direct comparisons of performance across architectures and initialization strategies. Python 3.11.0 was used to run all experiments, and the PyTorch 1.13.0 ^36^ library was used for DL. The “corrected resampled t-test” ^37,38^ was used to test for significant differences in model performance. The use of nested-cross validation violates the independence assumption of the traditional t-test ^39^, which could be used here.

## Results

### Segmentation

The segmentation results across 25 trials for all three DL architectures are shown in Figure 4. The MedNeXt segmentation model (mean Dice score: 0.555, 95% Confidence Interval (CI) for the mean: [0.529, 0.574]) outperformed both the CNN-based MedicalNet (0.516, [0.492, 0.537]) and the CNN-transformer hybrid TransBTS (0.449, [0.420, 0.482]). The differences between the performance of each of the models were statistically significant. The p-values of the corrected resampled t-tests when comparing the performance of the MedNeXt model to that of MedicalNet and TransBTS were 0.045 and 0.00014, respectively, while comparing the latter two models resulted in a p-value of 0.017.

**Figure 4:**
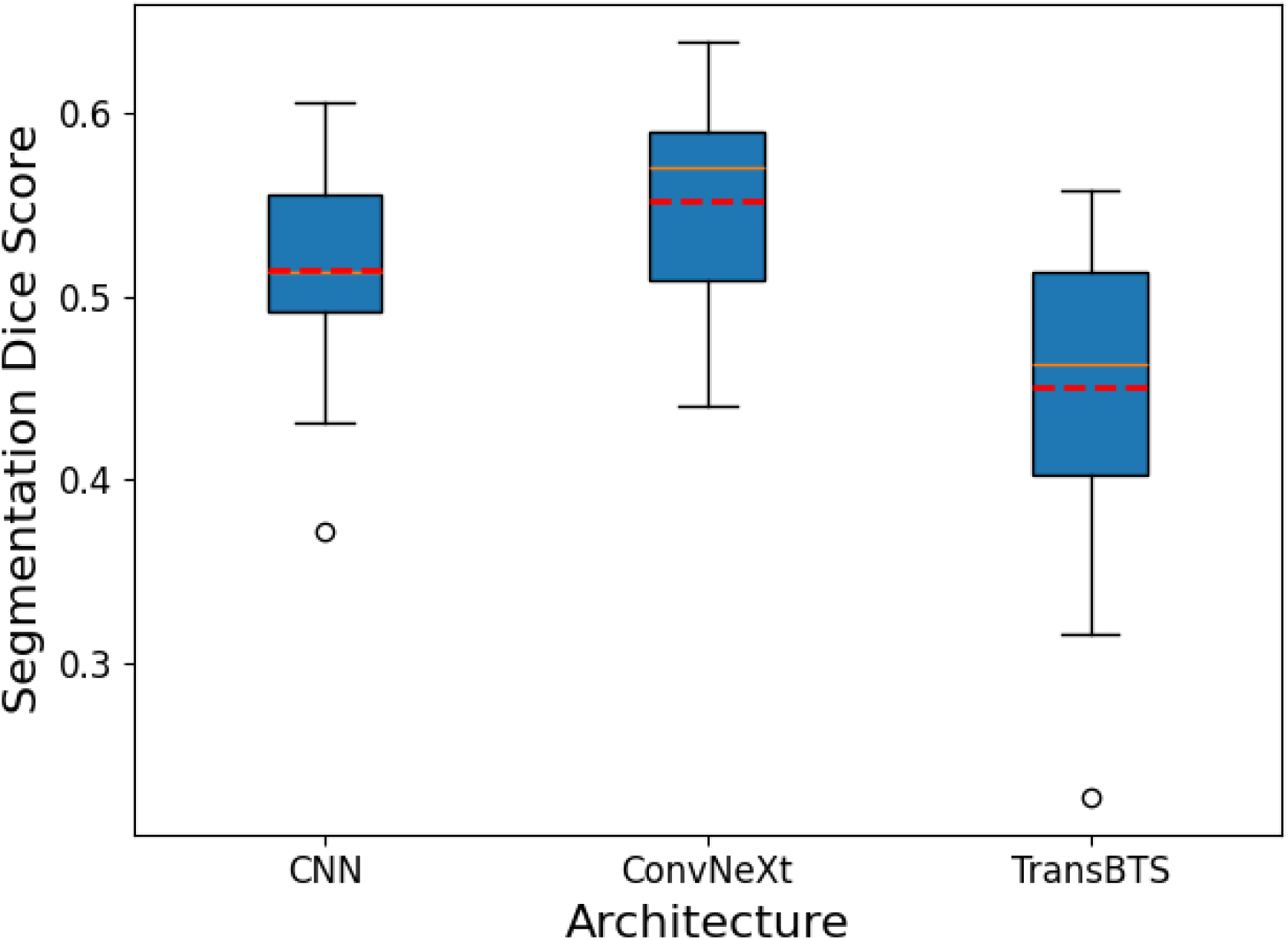
Distribution of AUC for three different DL architectures on the pLGG segmentation task across 25 trials. The solid orange line is the median, while the dashed red line is the mean.

### Classification

The mean three-class AUCs for all classification models are listed in Table 2. With weights initialized from scratch, the MedNeXt style classification model achieved a mean AUC of 0.747 (95% CI for the mean: [0.719, 0.774]) on the whole brain FLAIR MRI BRAF status classification task, where no segmentation was used. Performance improved (p-value: 0.0466) through pretraining on the segmentation, location, and radiomic feature value prediction tasks, resulting in a mean AUC of 0.779 (95% CI for the mean: [0.759, 0.799]), which was found to be the best classifier according to AUC. Additional performance metrics for this best model are listed in Table 3. Notably, despite a relatively high AUC, accuracy was determined to be relatively low overall (57%). The confusion matrix and per-class metrics in Table 3 suggest that this is because the model lacks the ability to differentiate between BRAF Mutation and non-BRAF altered tumors. Overall, the model is best at identifying BRAF Fusion: the AUC (0.867), precision (0.746), and recall (0.747) for this class were the highest.

**Table 2:** Comparison of the performance of different classification pipelines as measured by AUC. The best-performing pipeline, in bold, was the whole image model that was pretrained using the manual segmentation, radiomic features, and tumor location, before being fine-tuned for pLGG genetic status identification.

|  | **AUC**<br>**(95% Confidence Interval)** |
| --- | --- |
| **Manual Segmentation** | 0.756<br>(0.735, 0.778) |
| **Whole Image**<br>**(Random Initialization)** | 0.747<br>(0.719, 0.774) |
| **Whole Image**<br>**(Transfer Learning)** | 0.735<br>(0.704, 0.746) |
| **Whole Image**<br>**(Pretraining)** | **0.779**<br>**(0.759, 0.799)** |

**Table 3:**
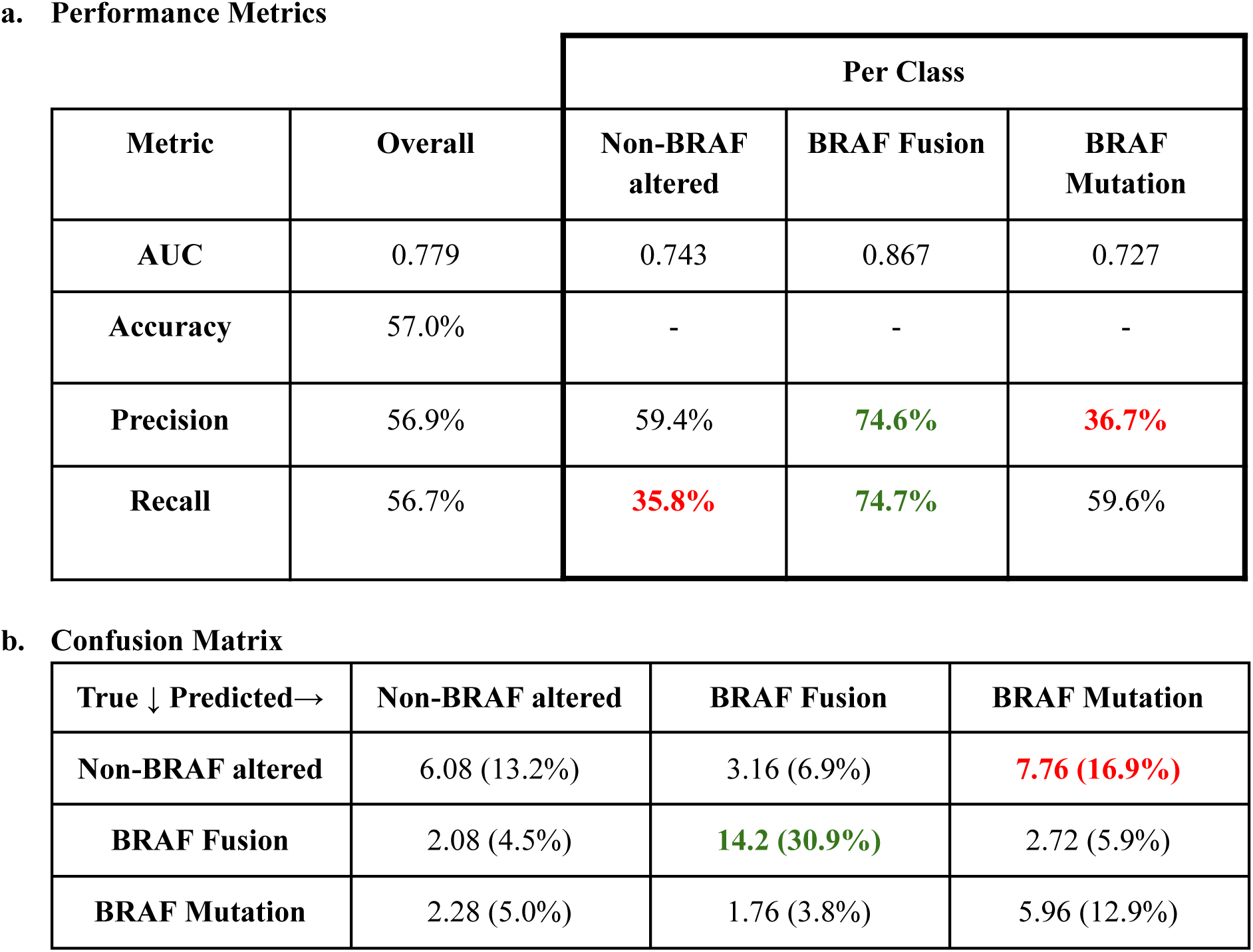
Detailed BRAF status classification performance metrics for model pretrained using manual segmentations, radiomics features, and tumor location. Throughout Table 3, values highlighted in green/red illustrate where the model is particularly strong (green) or weak (red). The first column of Table 3a describes the four metrics we used to evaluate the model. Each value in the table represents the mean of each metric, calculated across the 25 test sets. Accuracy was defined as the percentage of patients overall for which the true label was predicted by the model and was not calculated on a per-class basis. All other metrics were calculated according to their standard definition, for each class, and then those per-class values were averaged to obtain the values listed in the overall column. The second column of Table 3a details the values of the performance metrics overall. The remaining columns detail the performance metrics on a per-class basis. Table 3b contains the confusion matrix for the classification model, where the primary value is the mean of the number of patients in the cell across 25 trials, with the percentage of patients in parentheses. It can be seen from the confusion matrix that the model excels at identifying BRAF Fusion, but frequently mistakes non-BRAF altered tumors for BRAF mutations.

**a. Performance Metrics**
| Metric | Overall | Per Class |  |  |
| --- | --- | --- | --- | --- |
|  |  | Non-BRAF altered | BRAF Fusion | BRAF Mutation |
| AUC | 0.779 | 0.743 | 0.867 | 0.727 |
| Accuracy | 57.0% | - | - | - |
| Precision | 56.9% | 59.4% | 74.6% | 36.7% |
| Recall | 56.7% | 35.8% | 74.7% | 59.6% |

**Table 3:** Detailed BRAF status classification performance metrics for model pretrained using manual segmentations, radiomics features, and tumor location.
| True ↓ Predicted→ | Non-BRAF altered | BRAF Fusion | BRAF Mutation |
| --- | --- | --- | --- |
| Non-BRAF altered | 6.08 (13.2%) | 3.16 (6.9%) | 7.76 (16.9%) |
| BRAF Fusion | 2.08 (4.5%) | 14.2 (30.9%) | 2.72 (5.9%) |
| BRAF Mutation | 2.28 (5.0%) | 1.76 (3.8%) | 5.96 (12.9%) |

The baseline model, which used the tumor region alone as an input, performed similarly to the whole-brain FLAIR MRI models. The mean baseline AUC of 0.756 (95% CI for the mean: [0.735, 0.778]), was only slightly higher than the randomly initialized whole FLAIR MRI model, and the difference between the performance of the two models was not statistically significant (p-value: 0.342). The best-performing whole-brain FLAIR MRI model, pretrained on pLGG data, produced a higher AUC than the baseline, though again the difference was found to not be statistically significant (p-value: 0.141).

### Transfer Learning

TL from BraTS did not have a statistically significant impact on segmentation or classification performance. For the segmentation task, pretraining the MedNeXt style model on BraTS before fine-tuning it on our pLGG dataset resulted in a mean Dice score of 0.539 (95% CI for the mean: [0.525, 0.553]). This was lower than when training from scratch (0.555), and not significantly different (p-value: 0.180). Similarly, pretraining the MedNeXt classification model on BraTS prior to fine-tuning resulted in lower AUC (mean: 0.725, 95% CI for the mean: [0.704, 0.746]) than when training from scratch (0.747). Statistical testing showed that the difference was not significant (p-value: 0.215).

## Discussion

In this study, we introduced a novel, segmentation-free MRI-based pipeline to assess molecular status in patients with pLGG. Our whole-brain FLAIR MRI classification approach reduces the reliance of ML models used for pLGG genetic status classification on tumor segmentations. Our results illustrate that even without pretraining, and thus without any knowledge about tumor location whatsoever, the whole-brain FLAIR MRI classification model performs similarly to the baseline model that takes the tumor region as an input. Pretraining on pLGG tumor segmentations, and features derived from them, before fine-tuning on the classification task was found to improve model performance. This pretraining approach introduces a novel way of incorporating tumor information into a classification pipeline that does not rely on segmentations after training. The pretrained whole-brain FLAIR MRI classification model was able to accurately identify tumors with a BRAF Fusion but had difficulties differentiating between BRAF Mutations and non-BRAF altered tumors. These results suggest identification of BRAF Fusion, the most common genetic alteration in pLGGs, is the most promising use case for the whole-brain FLAIR MRI classification pipeline. We emphasize that once deployed, the pretrained model does not have any need for tumor segmentations.

We also explored the potential for advanced DL architectures to accurately and reliably segment MRIs of patients with pLGGs. Previous pLGG segmentation studies have relied on CNN-based models ^19,23^; we found that a modern architecture, based on the MedNext architecture, performed better. The MedNeXt model was designed using ConvNeXt ^27^ layers, which combine the inherent inductive biases of CNNs with the ability of transformers to scale efficiently and capture long-range dependencies ^32^. It appears that MedNeXt combines the best of CNNs and transformers; it was previously shown to outperform both CNNs and transformers for adult brain tumor segmentation ^32^. Our results suggest MedNeXt is better at pediatric brain tumor segmentation, too. However, for some patients, segmentations were poor (Figure 5), suggesting that automated segmentations are too unreliable for use in downstream tasks although they were sufficiently accurate to be helpful in pretraining. Manual segmentations are also suboptimal classification model inputs. They have known inter- and intra-rater reproducibility issues, and thus the classifier’s input and output can be different for the same case, harming clinician confidence in the model.

**Figure 5:**
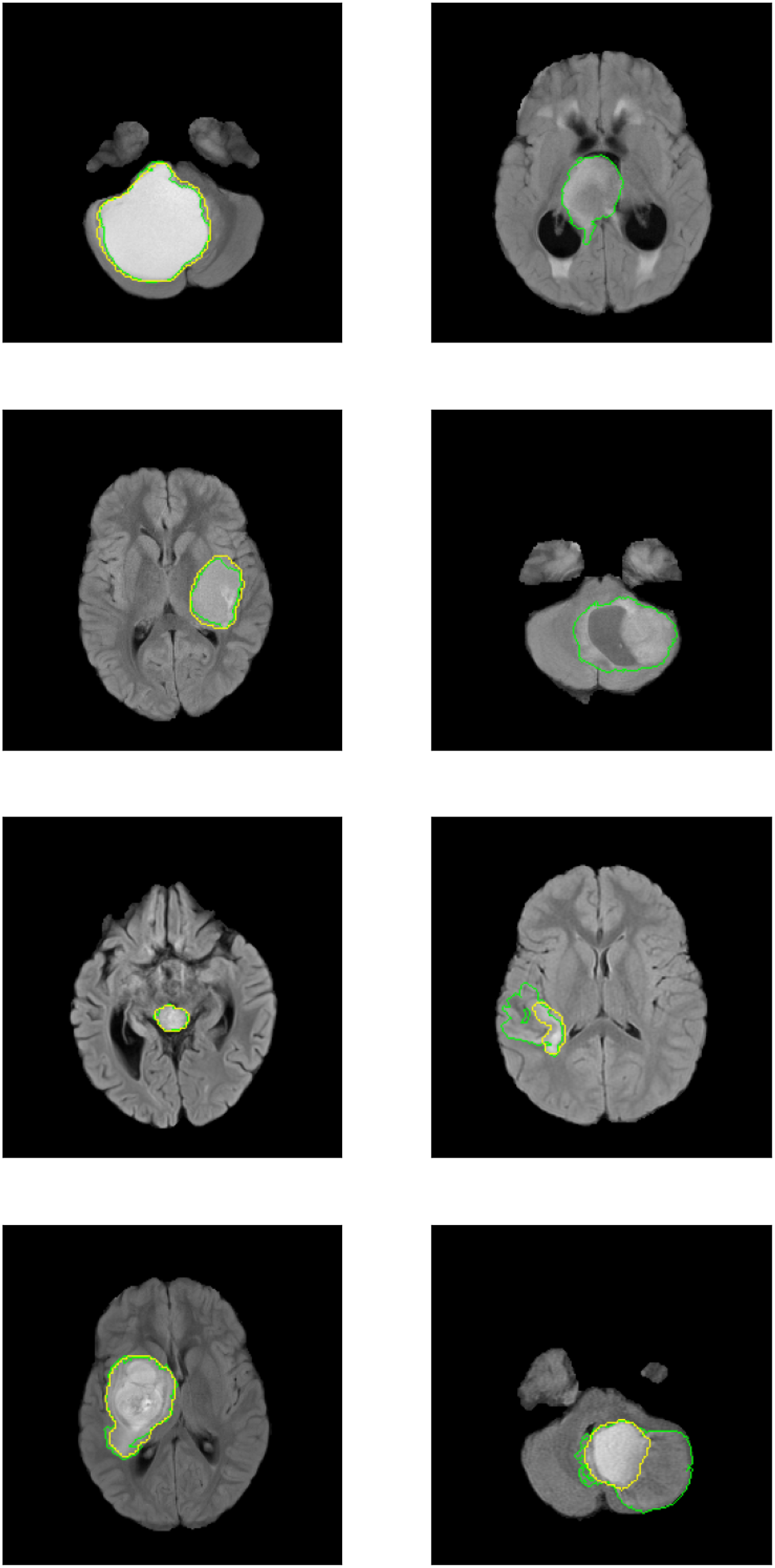
Preprocessed images for 8 patients, with both automated and manual segmentations. The yellow line delineates the predicted tumor region while the green area outlines the manually segmented tumor region. Similar to what has been found in previous works, our model works well for some patients (left) but fails in certain cases (right). Notably, for the two topmost cases in the right column, the segmentation is completely off, with no tumor identified at all in the slice depicted. A downstream classification model will struggle to identify the genetic status of a tumor accurately if the automated segmentation model it depends on does not correctly identify the tumor.

Notably, TL from BraTS did not improve classification or segmentation model performance. TL has successfully boosted model performance for numerous medical tasks ^33^. However, it has not always proven useful, likely due to domain shift, which is when there are differences between the dataset involved in pretraining and the dataset the model is later fine-tuned on ^40,41^. For example, as described in ^42^, previous studies focused on TL from BraTS for pLGG automatic tumor segmentation ^23,43^ did not find a consistent, substantial improvement in model performance using TL. Considering the significant differences between adult and pediatric brain tumors ^44–46^, it is perhaps unsurprising that knowledge does not transfer between the two patient groups. We hypothesize that our custom pretraining scheme involving manual segmentation, tumor location, and radiomic features, was successful because it did not rely on any additional data and thus was not susceptible to domain shift.

Our work is not without limitations. First, our observations about the relative performance of different DL models and training schemes are specific to the situations in which we evaluated them, namely, on our hospital’s pLGG dataset using a finite set of hyperparameters. Further tests on different tasks, datasets, and model configurations need to be run to determine the generalizability of these claims. However, the fact that similar results have been found for related tasks ^32^ supports our claims. Second, our model was best suited for identifying BRAF Fusion and had difficulties differentiating BRAF Mutations from non-BRAF altered tumors. This could potentially be addressed by data augmentation techniques such as synthetic image generation, which has proven successful in improving pLGG genetic status classification ^47^, but was not applied here. Furthermore, uncertainty quantification techniques such as evidential deep learning ^48^, or dropout as Bayesian approximation ^49^, could be applied to identify uncertain predictions, potentially facilitating the use of our model to distinguish between all 3 classes more accurately, or even to identify uncertain segmentations, enabling the use of automated segmentations for certain cases.

## Conclusion

We introduced the first model for pLGG genetic status assessment that does not rely on manual or automated tumor segmentations. Our training pipeline uses segmentations, and features derived from them, for pretraining, but once deployed, the final model does not require any segmentations to determine the genetic status of pLGG.

## Data Availability

The datasets generated and/or analyzed during the current study are available from the corresponding author on reasonable request pending the approval of the institution(s) and trial/study investigators who contributed to the dataset.

## List of Abbreviations

AUC: Area Under the Receiver Operating Characteristic Curve
BRAF Fusion: KIAA1549::BRAF fusion
BRAF Mutation: BRAF p.V600E mutation
CI: Confidence Interval
CNN: Convolutional Neural Network
CNS: Central Nervous System
DL: Deep Learning
FLAIR: Fluid-Attenuated Inversion Recovery
ML: Machine Learning
MRI: Magnetic Resonance Imaging
pLGG: Pediatric Low-Grade Glioma
TL: Transfer Learning

## Acknowledgments

This study was made possible by the financial support of the Canadian Institutes of Health Research (CIHR) (Funding Reference Number: 184015).

## Appendix

### DL architectures for brain tumor segmentation

CNN-based architectures, such as the U-Net models used in previous studies aiming to automate the segmentation of pLGGs ^19,23^, have built-in inductive biases ^50^. For example, most CNNs are to some degree translation invariant, depending on the extent to which pooling layers are included ^50,51^. Translation invariance means that these models tend to produce the same output when objects are shifted around within the input image. This can be a problem for the types of tasks we are considering here: the same signal on an MRI can have different meanings depending on which part of the brain it is located. As noted in ^52^, the lack of ability for CNNs to account for positional information limits their potential in medical image analysis tasks, such as tumor classification and segmentation, where anatomical location is a key feature.

The transformer architecture, which has revolutionized the field of natural language processing ^53^, has the potential to address the limitations of CNNs. In 2021, Dosovitskiy et al. introduced the vision transformer ^54^, adapting the original transformer model to the imaging domain. Vision transformers have fewer inductive biases than CNNs, making them more flexible ^50^. For example, unlike CNNs, the transformer architecture can capture long-range dependencies ^50,53^, making it capable of discovering features within an image that a CNN cannot due to its local receptive field. TransBTS ^26^, a hybrid model relying on convolutional layers to extract local features which are then used to encode global features through transformer-based self-attention layers, outperformed convolutional layers alone for adult brain MRI tumor segmentation. Similarly, Karimi et al. ^25^ showed that the performance of a convolution-free brain MRI segmentation model, relying solely on transformers, improved upon the results of CNN-based models.

Liu et al. ^27^ introduced the ConvNeXt architecture as another alternative to classic CNNs. Similar to a typical CNN, ConvNeXt uses only convolutional layers but it also incorporates design choices that are typically reserved for transformers, such as larger kernels, fewer activation functions, and Batch Normalization ^55^ instead of Layer Normalization ^56^. ConvNeXt was shown to be an efficient model, due to the inductive biases of the convolutional layers, while also maintaining the architectural improvements of transformers; it outperformed both traditional CNNs and transformers on a natural image classification task ^27^. Roy et al. ^32^ adapted the ConvNeXt architecture for medical image segmentation; their model MedNeXt, outperformed both purely CNN and transformer-based models, as well as hybrid approaches, for adult brain tumor segmentation, among other tasks.

More recently, there has been great interest in using generalist models like the Segment Anything Model (SAM) ^57^, or offshoots of SAM tailored for the medical space such as MedSAM ^58^, to automate the segmentation of brain tumors. However, these models are not completely automated, they require user prompts to help guide them toward the true tumor region. SAM and its derivatives are outside the scope of this study, which instead focuses on evaluating the performance of fully automated approaches for pLGG segmentation using the CNN, transformer, and MedNeXt architectures.

## Notes

### Competing Interest Statement

The authors have declared no competing interest.

### Funding Statement

This study was funded by the Canadian Institutes of Health Research (CIHR) (Funding Reference Number: 184015).

### Author Declarations

Research Ethics Board of of The Hospital For Sick Children (Toronto, Canada) waived ethical approval for this work.

